# Assessing the Relationship between Serum IGF-1 and BMI by Age in the Long Life Family Study

**DOI:** 10.1101/2020.06.30.20119743

**Authors:** Rehab A. Sherlala, Candace M. Kammerer, Allison L. Kuipers, Mary K. Wojczynski, Svetlana V. Ukraintseva, Mary F. Feitosa, Jonas Mengel-From, Joseph M. Zmuda, Ryan L. Minster

## Abstract

**Background:** Serum levels of insulin-like growth factor 1 (IGF-1) and body mass index (BMI) are both associated with susceptibility to age-related diseases. Reports on the correlation between them have been conflicting, with both positive to negative correlations reported. However, the age ranges of the participants varied widely among these studies.

**Methods:** Using data on 4,241 participants (aged 24–110) from the Long Life Family Study, we investigated the relationship between IGF-1 and BMI by age groups using regression analysis.

**Results:** When stratified by age quartile, the correlation between IGF-1 and BMI varied: in the 1st quartile (Q1, 24 y–58 y) the correlation was negative (*r* = −0.1, *P* = 0.0008); in Q2 (59 y– 66 y) it was negative (*r* = −0.035) but non-significant; in Q3 (67 y–86 y) it was positive (*r* = 0.045) but non-significant; and in Q4 (87 y–110 y) the correlation was positive (*r* = 0.14, *P* < 0.0001). This pattern did not differ by sex. We observed a similar age-related pattern between IGF-1 and BMI among participants in the third National Health and Nutritional Examination Survey.

**Conclusions:** Our results, that the relationship between IGF-1 and BMI differs by age, may explain some of the inconsistency in reports about their relationship and encourage additional studies to understand the mechanisms underlying it.

## Introduction

Insulin-like growth factor 1 (IGF-1) is a member of the IGF-1 pathway ^1^, which appears to play a key role in the processes underlying longevity ^2^. Many epidemiological studies report association of serum IGF-1 concentration with elevated risk of type 2 diabetes ^3^, cancer ^4^, cardiovascular disease ^5^, and mortality ^6^. IGF-1 has structural homology to insulin and characteristics of both a circulating hormone that mediates growth hormone (GH) actions in promoting growth, development, and metabolism ^1^ and a local tissue growth factor that promotes cellular growth, differentiation, and apoptosis ^7^. Serum IGF-1 levels are heritable, with estimated heritability ranging from 40% to 63% ^8,9^, and are influenced by obesity, age, sex, physical activity, GH level, and nutritional status ^9^. Across the lifespan, serum IGF-1 levels are low at birth, increase during childhood and puberty, and reach their highest concentration during early adulthood. They then start to decline in the third decade of life ^10^.

Body mass index (BMI) is also strongly associated with risk for chronic disease development associated with aging ^11^. Because both IGF-1 and BMI are associated with disease risk and disease endpoints, several studies have assessed the relationship between them. Understanding the relationship between these two predictors could help categorizing those at risk of disease development or event. However, the relationship between BMI and IGF-1 across studies is neither consistent nor clear. Several studies have reported that IGF-1 levels are inversely correlated with BMI ^12–17^; whereas others report a positive correlation ^18^ or no correlation ^19^. Most of these studies estimated the relationship between IGF-1 and BMI by stratifying the samples based on BMI categories, and not age, although the study participants’ ages varied widely ^14,16,17,19^.

To date, no study has assessed the influence of age on the association between BMI and IGF-1. We hypothesize that the relationship between IGF-1 and BMI varies by age.

Here we present a cross-sectional study of the relationship between IGF-1 and BMI in a large sample of 4,241 participants from the Long Life Family Study (LLFS) with validation of the relationships in a large sample of 2,555 participants from the third National Health and Nutritional Examination Survey (NHANES III). In particular, we assessed whether the relationship between IGF-1 and BMI varies in an age- and sex-specific manner.

## Methods

### Study population

The primary sample for this study is a set of participants from LLFS. LLFS is a multi-center family-based cohort study of 539 families that was designed to determine the genetic and behavioral/environmental risk factors that promote exceptional longevity ^20^. The families were recruited between 2006 and 2009 from the USA and Denmark at four enrollment sites (New York, Boston, and Pittsburgh in the United States and nationwide in Denmark). More details of the LLFS cohort are described in ^20^. The total number of enrolled participants is 4,953, consisting of long-lived probands and their siblings (*n* = 1,727), the offspring of this generation and their spouses (*n* = 3,226). Participants without measurements of serum IGF-1 levels or BMI were excluded, therefore, the total sample size for this analysis is 4,241 participants (aged 24 y–110 y) consisting of 1,391 from the proband generation (49 y–110 y), 2,119 from the offspring generation (32 y–87 y), and 731 offspring spouses (24 y–88 y). All participants self-identified as non-Hispanic White.

The findings in LLFS were replicated using participants from NHANES III ^21^. NHANES III is a probability sample of ∼39,000 participants aged 2 months and older and was designed to be representative of the US population. It was conducted from 1988 to 1994 in two phases. Of the total sample of adults (*n* = 20,024), we selected a subset of 2,555 non-Hispanic White participants (20 y–90 y) with complete record of the study variables. Parallel analyses were conducted with non-Hispanic Black participants (*n* = 1639, 20 y–90 y) and among Mexican American participants (*n* = 1607, 20 y–90 y).

### Anthropometry

In both LLFS and NHANES III, standing height, weight, and waist circumference (WC) were assessed by trained interviewers with a standardized protocol and skill level. BMI was calculated as weight in kilograms per the square of the height in meters. Age, race, ethnicity, and sex were taken by self-report during the interview.

### Laboratory assays

In LLFS, fasting peripheral blood samples were obtained from participants and then shipped to the Advanced Diagnostics and Research Laboratory at the University of Minnesota ^22^. IGF-1 was measured in serum using a solid-phase enzyme-linked chemiluminescent immunoassay on an Immulite 2000 system (Siemens Healthcare Diagnostics, Inc.). The inter-assay coefficient of variability was 8.7%.

In NHANES III, fasting serum samples were collected from 1988 to 1994 and IGF-1 concentrations were quantified by IGF-I enzyme-linked immunosorbent assay (DSL 10-5600) including an extraction step which separates IGF-1 from its binding protein. The samples were reanalyzed if the coefficient of variation for replicate samples was greater than 15% ^23^. In the current study, we did not compare the absolute value from each type of assay.

### Statistical Approach

All data analysis was performed using R version 3.4.0 ^24^. To approximate normality, both IGF-1 and BMI were natural log–transformed for the analysis. We calculated the Pearson coefficient of correlation between IGF-1 levels and BMI, height, weight, waist circumference, and age.

We used two sample t-tests to assess the mean age difference and mean IGF-1 difference between the LLFS and NHANES III. Also, we used the *lstrends* function to estimate and compare the slopes of fitted lines between male and females in both studies.

We regressed log(IGF-1) on log(BMI) to get an overall assessment of their relationship in each sample. We performed linear mixed effect model using *coxme* package ^25^ and adjusting for covariates. Covariates were chosen based on their known association with serum IGF-1 and included: age, age^2^, male sex, field center, log(BMI) × age, and log(BMI) × sex as fixed variables, and also adjusting for kinship as a random variable to account for the relatedness between LLFS participants. To assess whether the relationship differs by sex, for all analyses described below we also stratified by sex, and regressed IGF-1 on BMI as above without the sex and log(BMI) × sex terms.

First, we regressed IGF-1 on BMI in all samples regardless of age. Then, to assess the relationship between IGF-1 and BMI by age, we divided the LLFS sample into age quartiles and performed linear mixed-model regression of IGF-1 with BMI as a fixed variable and kinship as a random variable within each age quartile.

In addition, we also conducted all of the previous analyses with IGF-1 and WC, as another measure of adiposity.

To validate the results, the same approach was used with the NHANES III sample of 2,555 non-Hispanic White participants, although we did not adjust for kinship, as the participants were assumed to be unrelated. The age-quartile thresholds in LLFS were used as age-group thresholds in NHANES III. We then assessed whether the relationship between IGF-1 and BMI also differed by age quartile in non-Hispanic Black (*n* = 1,639) and Mexican American (*n* = 1,607) participants.

## Results

The means, standard deviations and proportions of key characteristics of the study samples are presented in **Table 1**. The Long Life Family Study participants’ mean age was 70 years with a range of 24 years–110 years. The overall mean serum IGF-1 level was 128.3 ng/mL and ranged from 26 ng/mL to 745 ng/mL. The mean BMI was 27 kg/m^2^. In NHANES III, the participants’ mean age was 53.2 years with a range of 20 years–90 years. The overall mean IGF-1 was 249.5 ng/mL and ranged from 25.3 ng/mL to 863.8 ng/mL. The mean BMI was 26.48 kg/m^2^.

**Table 1.** Descriptive statistics of age, IGF-1, and anthropometric measurements in LLFS and NHANES III.

| **Measurements** | **LLFS** |  |  | **NHANES III** |  |  |
| --- | --- | --- | --- | --- | --- | --- |
|  | **Male (45.3%)** | **Female (54.7%)** | **Total** | **Male (45.2%)** | **Female (54.8%)** | **Total** |
| Participants (*n*) | 1924 | 2317 | 4241 | 1155 | 1400 | 2555 |
| 1<sup>st</sup> Age Quartile/Group (*n*)<br>(20 y–58 y) | 421 | 640 | 1061 | 643 | 840 | 1483 |
| 2<sup>nd</sup> Age Quartile/Group (*n*)<br>(58 y–66 y) | 507 | 553 | 1060 | 140 | 144 | 284 |
| 3<sup>rd</sup> Age Quartile/Group (*n*)<br>(66 y–86 y) | 497 | 563 | 1060 | 346 | 381 | 727 |
| 4<sup>th</sup> Age Quartile/Group (*n*)<br>(87 y–110 y) | 499 | 561 | 1060 | 26 | 35 | 61 |
|  | **Mean (s.d.)** | **mean(s.d.)** | **mean(s.d.)** | **mean(s.d.)** | **mean(s.d.)** | **mean(s.d.)** |
| Age at Enrollment (y) | 70.6 (15.3) | 69.5 (15.9) | 70.0 (15.6) | 54.0 (19.5) | 52.6 (19.7) | 53.2 (19.6) |
| IGF-1 (ng/ml) | 134.6 (54.0) | 123.2 (51.6) | 128.3 (52.9) | 264.7 (97.6) | 236.9 (103.6) | 249.5 (101.4) |
| BMI (kg/m<sup>2</sup>) | 27.5 (4.0) | 26.7 (5.3) | 27.8 (4.8) | 26.7 (4.8) | 26.3 (5.7) | 26.5 (5.3) |
| Height (cm) | 173.6 (7.7) | 159.6 (7.8) | 166.0 (10.5) | 175.5 (7.1) | 161.3 (7.0) | 167.7 (9.9) |
| Weight (kg) | 83.2 (14.9) | 68.2 (14.9) | 75.0 (16.5) | 82.5 (16.5) | 68.4 (15.4) | 74.8 (17.4) |
| WC (cm) | 99.5 (11.0) | 90.4 (13.9) | 94.5 (13.4) | 97.7 (12.7) | 89.5 (14.5) | 93.2 (14.3) |

*Note*, enrollment for LLFS was 2006–2008, and for NHANES III was 1988–1994. IGF-1 and
BMI are values on the natural scale to report the mean and standard deviation. BMI = body mass
index, IGF-1= insulin-like growth factor, WC= waist circumference.

Both studies had a wide age range, but on average, NHANES III participants were 16.8 years younger than LLFS participants (*p* < 0.0001). The age distribution in LLFS is bimodal due to the family-study design with some overlap between the LLFS generations; the age distribution in NHANES III is approximately uniform across its range (**Appendix Figure 1**). As expected, log(IGF-1) levels were negatively correlated with age in both LLFS and NHANES III, *r* = −0.418 (*P* < 0.001) and *r* = −0.475 (*P* < 0.001), respectively **(Figure 1a**). In addition, mean serum IGF-1 levels were 121.3 ng/ml lower in LLFS compared to NHANES III (*p* < 0.0001).

**Figure 1.**
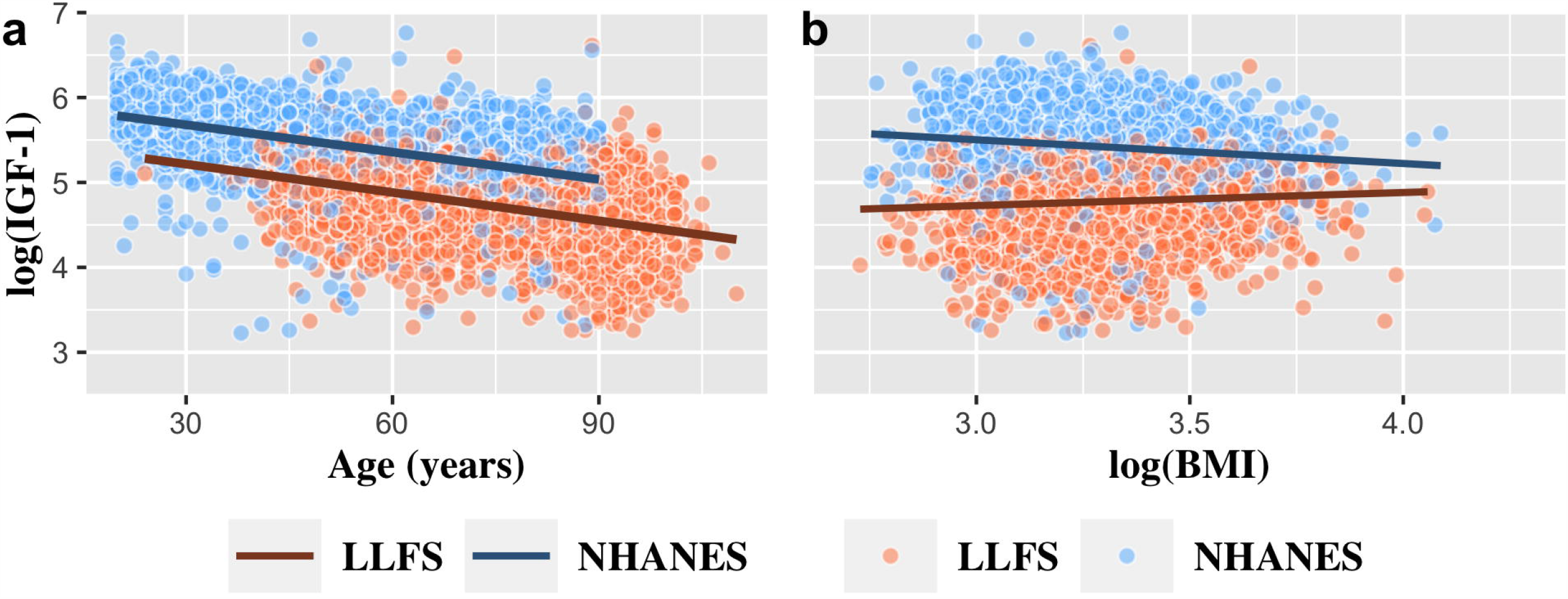
**a, b) a**. Scatter plot of log (IGF-1) by age and **b**. Scatter plot of log(IGF-1) by log(BMI) in both, LLFS and NHANES III

In LLFS, across all participants, log(IGF-1) levels were positively correlated with both log(BMI) and WC, *r* = 0.062 (*P* < 0.001) and *r* = 0.032 (*P =* 0.06), respectively (**Appendix Table 1.1**). In contrast, in NHANES III, log(IGF-1) levels were negatively correlated with log(BMI) and WC, *r* = −0.119 (*P* < 0.001) and *r =* −0.175 (*P* < 0.001), respectively. However, log(IGF-1) was positively correlated with height among both LLFS and NHANES III participants (*r* = 0.27, *P* < 0.001 and *r* = 0.24, *P* < 0.001, respectively) (**Appendix Table 1.2**).

In the regression analysis adjusted only for the kinship in LLFS, log(IGF-1) was associated positively with log(BMI) (β = 0.164, *P =* 1.6 × 10^−5^); whereas in NHANES III, the relationship was significant and negative (β = −0.3, *P =* 1.4 × 10^−9^) (**Table 2, Figure 1b)**.

**Table 2.**
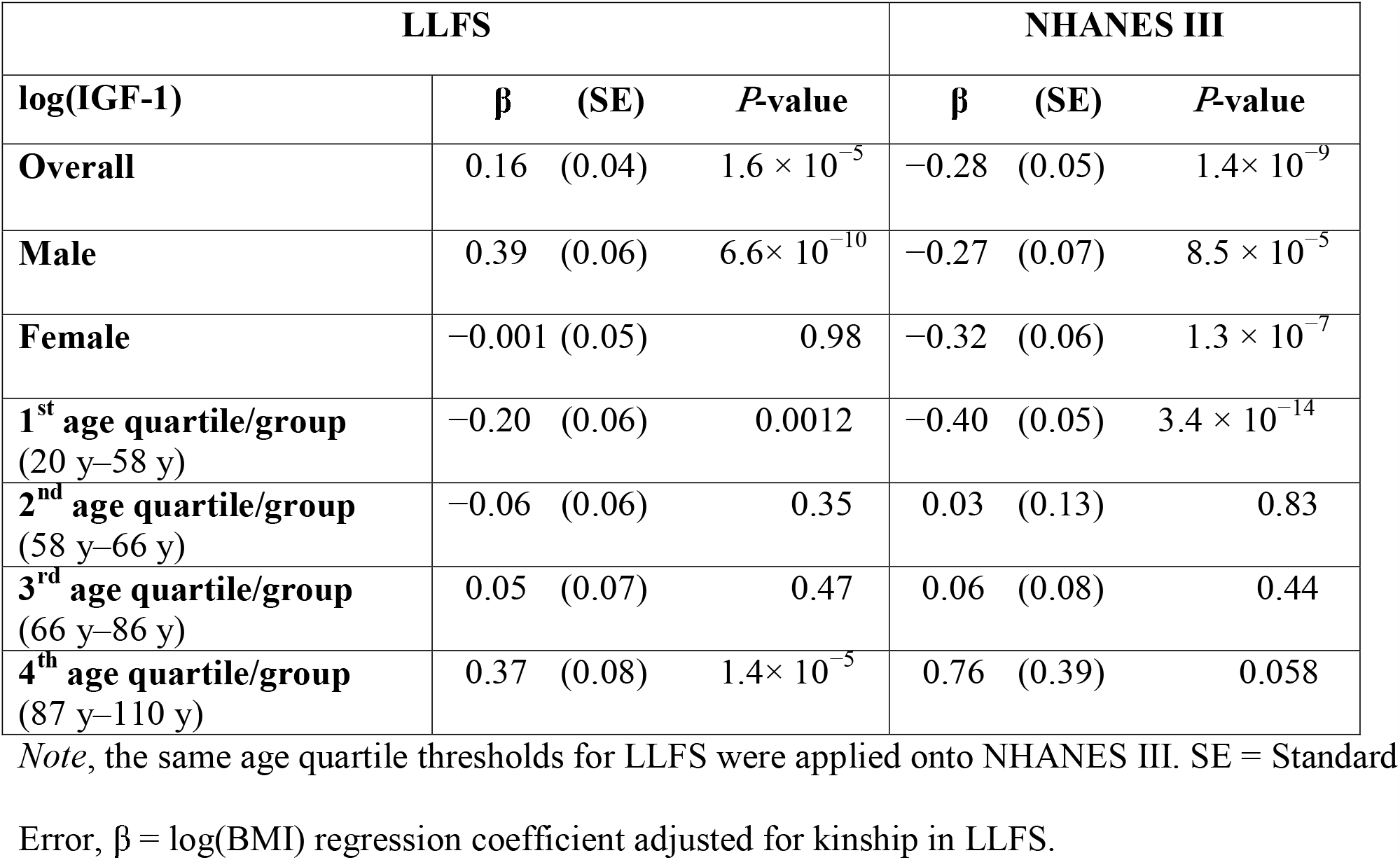
Result of linear regression of association between log(IGF-1) as an outcome and log(BMI) in the overall sample and per gender of LLFS and NHANES III and then stratified by age.

In LLFS, interaction for both log(BMI) and age, and log(BMI) and sex had significant effects on log(IGF-1) (both *P =* <0.0001) **(Appendix Table 2)**. Whereas in NHANES III, there was a significant interaction effect between log(BMI) and age (*P =* 5.3 × 10^−5^) on log(IGF-1), but no significant interaction between log(BMI) and sex (*P* = 0.5) (**Appendix Table 2**).

We further investigated the interaction between age and log(BMI) on log(IGF-1) in LLFS using age quartiles (**Appendix Table 3)**. As can be seen in **Figure 2a**, the relationship between log(IGF-1) and log(BMI) differed by age quartile. In the first (youngest) age quartile (20 y–58 y) the relationship was significant and negative (β = −0.2, *P =* 0.0007), in the second (59 y–66 y) and third (67 y–86 y) age quartiles the relationship was non-significant, but in the fourth (oldest) quartile (87 y–110 y), the relationship was significant and positive (β = 0.39, *P =* 5.3 × 10^−6^) (**Table 2**). When the NHANES III data was stratified using the LLFS age-quartile thresholds, a similar pattern was observed (**Table 2, Figure 2a**). We also stratified LLFS and NHANES III using age thresholds derived from NHANES III and applied them to LLFS; a similar pattern was observed (**Appendix Table 4, Appendix Figure 2**).

**Figure 2.**
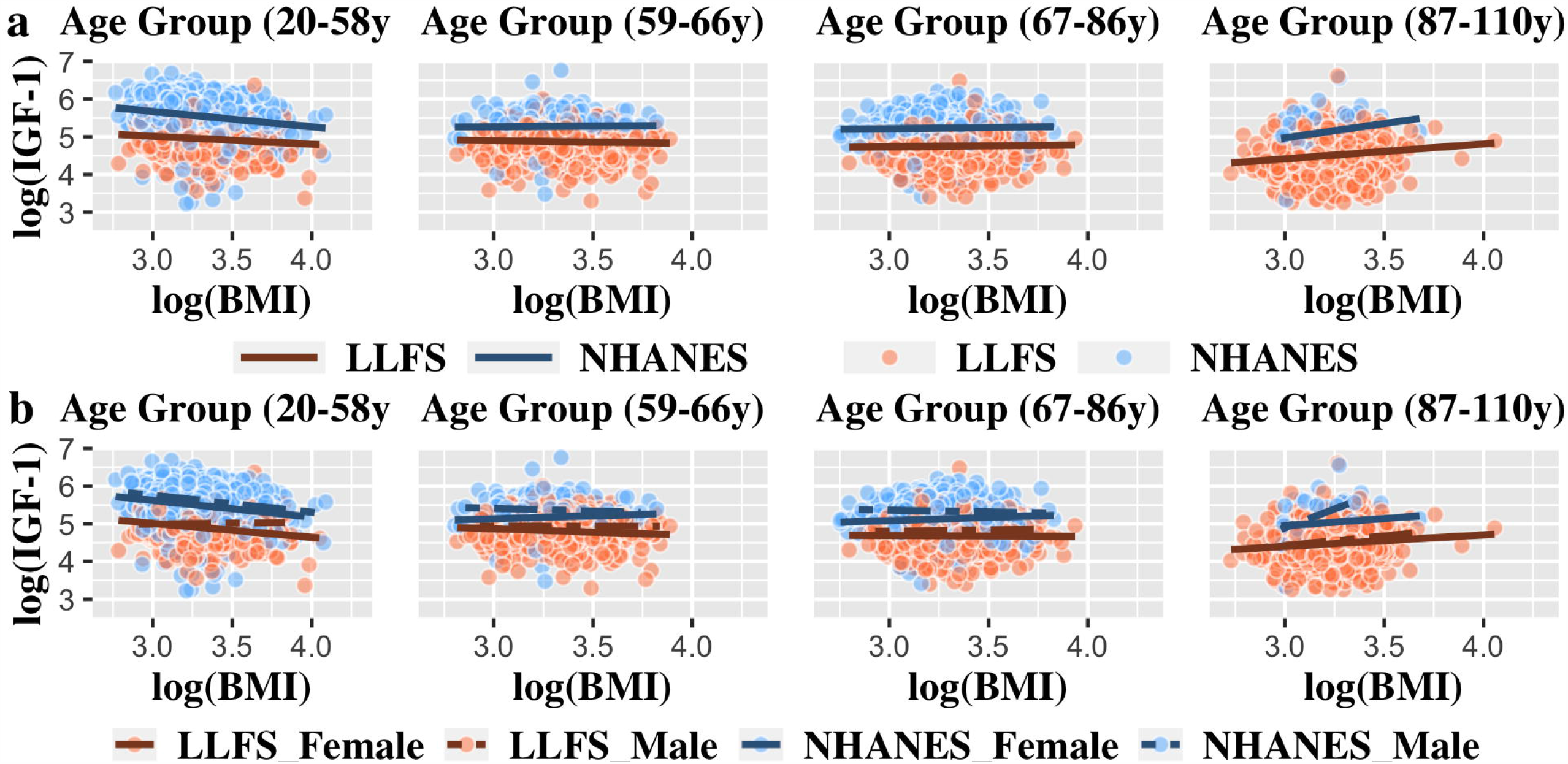
**a)** Scatter plot of log(IGF-1) by log(BMI) stratified by age groups, **b)** scatter plot of log(IGF-1) by log(BMI) per age groups and stratified by gender in LLFS and NHANES III. The same age quartile thresholds for LLFS were applied onto NHANES III

We next investigated the relationship between log(BMI) and sex on log(IGF-1) levels in LLFS by stratifying each age quartile by sex. Among females, log(IGF-1) was significantly and negatively associated with log(BMI) in the first (20 y–58 y) and second (59 y–66 y) age quartiles (β = −0.37, *P* = 5.5 × 10^−7^ and β = −0.17, *P* = 0.03, respectively), but there was no significant association in male (**Appendix Table 5, Figure 2b**). In the third age quartile (67 y–86 y), log(IGF-1) was not associated with log(BMI) in either sex. However, in the oldest quartile (87 y–110 y), log(IGF-1) was significantly and positively associated with log(BMI) in both sexes (β = 0.5, *P* = 0.003 and β = 0.3, *P* = 0.006, respectively) (**Appendix Table 5, Figure 2b**). The relationship between log(IGF-1) and log(BMI) did not significantly differ by sex in NHANES III, except in the fourth quartile (87 y–110 y) wherein the association was positive and significant in male (*P* = 0.01), but not in female (*P* = 0.4), though the sample sizes were small (**Appendix Table 5, Figure 2b**). In addition, we observed a significant slope difference by sex in the relationship between log(IGF-1) and log(BMI) in the first age quartile of LLFS only (*P* = 0.002)

Similar results were observed for the relationship between IGF-1 and WC in both studies (**Appendix Table 6, Appendix Figure 3**). Also, similar patterns were seen among non-Hispanic Black participants and among Mexican American participants in NHANES III (**Appendix Table 7, Appendix Figure 4)**.

## Discussion

In this cross-sectional study of LLFS, a unique family-based cohort of exceptional longevity, we examined the age- and sex specific effects of the relationship between serum IGF-1 levels and BMI. Younger participants (24 y–58 y), had a negative relationship between IGF-1 and BMI, while older participants (87 y–110 y) had a positive relationship. There was no statistically significant relationship for the age-groups in between. The same pattern was observed in an independent sample of non-Hispanic White adults of similar age range recruited from the general population in the NHANES III. In addition, we did not observe a consistent sex-specific difference in the relationship between IGF-1 and BMI across the age groups. The discrepancies in the relationship between serum IGF-1 and BMI among studies in the literature ^12–19^, may be explained by the ages of the cohorts used in the previous studies. Studies reporting the negative relationship between BMI and IGF-1were primarily conducted in participants with ages ranging from 10 years to 60 years ^12–17^. In contrast, studies that reported a positive relationship were often performed with older individuals with ages ranging from 45 years to 90 years^18^. These studies also showed similar pattern between the relationship of IGF-1 and WC ^14–16,18^. In addition, several of these studies were comprised of highly selected groups, such as obese/ overweight individuals, who might be experiencing weight-related disruption in insulin and growth hormone secretion ^16^.

The age-related difference in the relationship between IGF-1 and BMI might be due to height, which is a component of BMI. However, the pattern between IGF-1 and WC (a measure of central adiposity that is independent of height) was similar to that with BMI. This result indicates that the relationship is driven by adiposity rather than height (**Appendix Figure 3**).

In non-Hispanic Black and Mexican American participants, other investigators have reported an inverse association between IGF-1 and BMI ^12,14^; whereas studies in African Americans have reported no association ^27^. However, in our study, we saw similar patterns by age group in non-Hispanic Black and Mexican American participants within NHANES III as we saw in the non-Hispanic White participants, despite smaller sample sizes. (**Appendix Figure 4**). These results suggest that the relationship between IGF-1 and BMI by age group is similar among different racial/ethnic groups.

In this cohort we observed that younger participants had higher mean IGF-1 level compared to older participants, and this is consistent with known IGF-1 biology in adolescent and early-adulthood ^10^. Although the interaction between log(BMI) and sex in predicting log(IGF-1) was a statistically significant, the slope difference between men and women was not statistically significant except in the youngest LLFS age quartile. The latter results might reflect the sex differences in development during puberty and early adulthood.

LLFS and NHANES III data were collected 10 years apart, thus, period or cohort effects may exist, in addition to the age-effect we demonstrate. However, despite this potential period effect, the patterns were consistent for both LLFS and NHANES III cohorts, for BMI and WC, and across different racial/ethnic groups. In addition, mean serum IGF-1 levels were 121.2 ng/ml lower in LLFS compared to NHANES III. The most likely reason for this difference is the use of different assay kits in the measurement of serum IGF-1 levels between the two studies. Previous studies have reported significant differences in serum IGF-1 levels when using different assay kits, even though the samples were from the same population ^28–30^. Different assay kits have different age- and sex-specific reference ranges, and this might affect the upper and lower limit of each study’s serum IGF-1 levels. Furthermore, NHANES III was conducted using a sample from the general population, whereas LLFS sampled healthy long-lived individuals. Thus, study population and assay type are confounded, and it is impossible to determine if the difference in mean IGF-1 levels between the cohorts is due to ascertainment differences or to assay differences given these data. However, all statistical comparisons in this paper were done within-study, so the mean differences in IGF-1 levels between studies should not affect our conclusions, especially given that the patterns across age groups were similar.

This current study was cross-sectional and not longitudinal, therefore we could not measure the relationship between IGF-1 and BMI on the same participants throughout their lifespan in order to determine the patterns of change in this relationship. Instead we stratified our samples by age group and are extrapolating these cross-sectional results to reflect individual changes related to aging. However, additional longitudinal data are needed to confirm these findings. Also, we relied on BMI and WC to measure adiposity, which capture body size but not body composition. These anthropometric measurements are not as precise as imaging-based measurements, such as fat mass as estimated from dual X-ray absorptiometry or peripheral quantitative computed tomography ^31^. Such measures were not available for these studies. Another limitation in our study is the lack of data on potential confounders such as physical activity, diet, and GH level. These factors have major effects on BMI and IGF-1 level, and might also affect the result.

In summary, we identified age-related differences in the relationship between serum IGF-1 levels and BMI, as well as WC, in non-Hispanic White, non-Hispanic Black, and Mexican American participants. Further investigation into the underlying biology affecting the relationship between serum IGF-1 and measures of adiposity across the lifespan is needed to understand these observations. Such an understanding might help categorize individuals at risk of disease or inform interventions to delay disease depending on their age-dependent BMI and IGF-1 level.

## Data Availability

the data element will be included in this manuscript is not available online.

## Acknowledgments

We would like to thank the LLFS and NHANES III participants as well as the research staff who recruited and survey them.

